# Examining the psychometric properties of a crisis recovery in psychosis (CRISP) measure

**DOI:** 10.1101/2025.10.27.25338860

**Authors:** Lisa Wood, Hazel Chan, Rhys Thomas

**Affiliations:** Research and Development, North East London NHS Foundation Trust, Goodmayes Hospital, barley Lane, IG 3 8XJ; Division of Psychiatry, University College London, 149 Tottenham Court Road, London W1T 7NF

## Abstract

**Background:** Psychotic crises are complex idiosyncratic experiences and currently there are no validated measure which examine the recovery from such experiences. The aim of this study was to develop and psychometrically validate the crisis recovery in psychosis (CRISP) measure.

**Methods:** The 23-item CRISP was developed in partnership with a coproduction group of people with lived experience, family/carers and clinicians who had experience of mental health crisis care. A longitudinal design was adopted with 80 participants participating in the study, completing the CRISP along with other relevant measure at three time points to psychometrically validate it. The CRISP was examined for internal consistency, test-retest reliability, criterion validity, construct validity, sensitivity to change and floor and ceiling effects.

**Results:** Initial examination of item performance resulted in 9 items being removed due to low correlations (<0.2) with other items. A principal components varimax rotation produced a 3-factor solution, which included 14-items. The measure was demonstrated to be internally reliable (Cronbach alpha= 0.912 (CI: 0.880 - 0.938, p <0.001)) and reproducible (Cronbach alpha= 0.842 (CI: 0.742 to 0.904,p<0.001). The measure was also deemed to be valid and demonstrated criterion validity, construct validity, and sensitivity to change. No floor or ceiling effects were identified.

**Conclusion:** The CRISP is a simple, reliable, and valid measure of psychotic crisis recovery that measure core outcomes relevant to inpatient/crisis populations. Future research needs to be undertaken to examine its interpretability.

## Introduction

People experiencing psychosis often hold unusual beliefs and may see or hear things that other cannot (British Psychological Society, 2014). Over 1% of the population have such experiences and they usually begin between the ages of 14 – 35 (National Institute of Health and Care Excellence, 2014). For various reasons, such as trauma, stressful life events, or sudden stopping of anti-psychotic medication, experiences of psychosis can often escalate into a crisis (Wood et al., 2020). (L Wood et al., 2019). People experiencing psychosis make up to two third of those experiencing a mental health crisis (NHS Digital, 2020). When people with psychosis are in crisis, they are likely to receive care from crisis mental health services, such as psychiatric inpatient care, and crisis resolution home treatment teams. These services aim to contain a crisis, reduce risk, for example, self-harm and suicide, and improve an individual’s quality of life (Johnson, 2013).

A crisis has been defined as an acute grief reaction which is short lived that can derail an individual from their life goals (Black & Flynn, 2021). A crisis develops, and is perpetuated by, the individual’s perception of the event as extremely distressing and upsetting, alongside an inability to resolve the crisis through normal coping mechanisms (Roberts & Ottens, 2005). A crisis will cause disruption to a person’s daily functioning, cause acute distress, and put people at risk to themselves or others (for example, through self-harm and suicide) (Tobitt & Kamboj, 2011). A psychotic crisis is characterised by key psychological components such as a lack of control, risk of harm, functional disruptions and extreme emotional distress (Tobitt & Kamboj, 2011). It can also cause distressing psychotic symptoms such as command hallucinations and persecutory delusions.

Outcome measures are tools which can be used to describe the progress of care, support and treatment. Patient reported outcome measures are defined as information about the status of a patient’s health condition that comes directly from the patient, without interpretation of the patient’s response by a clinician or anyone else (NHS Digital, 2025). Outcome measures examining patient recovery have been criticised for being too symptom focused, and not focusing enough on the patient’s quality of life (Neil et al., 2009). This is particularly pertinent when a patient is in crisis as outcome measures usually examine risk minimisation and symptom reduction (Department of Health, 2024). Indeed, these clinical measures are important indicators of a crisis improving however there are several other key outcomes that just as important from a patient perspective. Service user led recovery research has identified that other outcomes that relate to rebuilding self, rebuilding life and hope for a better future are just as important as symptom and risk outcomes, if not more important (Pitt et al., 2007). There have been measures developed for community contexts to measure personal recovery, such as the process of recovery questionnaire (QPR) (Law & Morrison, 2014). However, no such measures have been developed for crisis settings where recovery outcomes are arguably different. Given the threshold for admission relates to the presentation of acute symptoms and high levels of risk, recovery from this setting is likely to include a combination of both person and clinical recovery factors.

Core outcome sets are now best-practice for researchers to bring a standardised approach to measuring outcomes of treatments or interventions (Chevance et al., 2020). For inpatient settings, a recent study has demonstrated that psychological interventions conducted in these settings should be measuring the outcomes of ability to Cope, Hopefulness, Quality of Life, Psychosis Symptoms, Mood, and Self-Harm Behaviours (Jacobsen et al., 2024). However, there are very few patient reported outcome measures that are psychometrically validated for use in crisis settings that measure crisis related outcomes.

There is a clear need to develop and psychometrically validate and patient reported outcome measure that can be used to measure recovery from a mental health crisis to ensure that patient outcomes can be prioritised in this context. The aim of this study was to develop and psychometrically validate an outcome measure examining key factors of recovery from a psychotic crisis.

## Methodology

### Design

Full Health Research Authority (HRA) and NHS Research Ethics Committee (REC) approval was granted (IRAS ID: 272043; 20/LO/0137/AM01) and the study was sponsored by the University College London. This study was a longitudinal study which was conducted across three time points to examine the reliability and validity of the CRISP. This study has followed guidance from Terwee et al (2007) in the development and psychometric validation of the measure. This study examined various forms of reliability and validity. Their definition and criteria that we adopted can be found in table 1.

### Development of the measure

Item generation for the CRISP was derived from several sources: a systematic review of recovery from psychosis within the inpatient setting (Wood & Alsawy, 2016), a systematic review of psychological interventions for psychosis in inpatient settings (Wood et al., 2020), and a series of qualitative interviews conducted with stakeholders about their priorities (Wood et al., 2018; L. Wood et al., 2019; L Wood et al., 2019). Content analysis was undertaken and codes were extracted from the data and collated (Krippendorff, 2004). For the qualitative data the full results section was used as data (both author interpretation and participant quotes) and for the quantitative studies outcome measures were screened and outcomes relevant to crisis recovery were extracted. These were either individual items on outcome measures or a type of outcome (e.g. readmission). The codes were then categorised into key areas of potential interest.

### Service user involvement

The item categories were presented to a coproduction group of eight people with lived experience of psychosis and inpatient care, family/carers, and clinicians. More detail about this group can be found here (Wood et al., 2023). The group then discussed the categories and decided on key items that were important for inclusion. The also gave feedback on the layout of the measure, design of the measure (i.e. clinician rate or self-report), scoring of the measure, and instructions.

### Final measure and scoring

The final measure was a 23-item self-report measure examining key crisis recovery items. It was rated on a scale from 1 (not at all) to 5 (all of the time). The items fell into one of 10 areas of interest comprising risk behaviours, emotional distress, symptoms of psychosis, physical health, cognitive changes, relationships, functioning, social problems, coping with crisis, and help seeking. The measure examined the person’s experience with these areas in the last week. The measure was self-report but could be completed with support from a family member, carer or mental health professional.

### Participants

Participants were recruited from two sources. Participants were either recruited from (a) an adapted Cognitive Behaviour Therapy for psychosis (CBTp) trial for inpatient settings trial (the CRISIS study, (Wood et al., 2022)) or (b) an outer London acute psychiatric inpatient hospital. In both (a) and (b) potential participants were identified by ward staff on participating wards who shared information about the study. If interested, they were then approached by a research assistant to take part. Inclusion criteria for participants will be (i) aged 18 and above; (ii) who meet criteria for a schizophrenia-spectrum diagnoses (schizophrenia, schizophreniform disorder, schizoaffective disorder, delusional disorder or psychotic disorder not otherwise specified; ICD-10), or meet criteria for an early intervention service (EIS) for treatment of psychosis to allow for diagnostic uncertainty; (iii) able to give informed consent and have the capacity to consent; (iv) currently receiving care from an acute psychiatric inpatient team; and (v) able to complete the research in English.

Exclusion criteria will be (i) non-English speakers (due to translation costs and difficulty of producing valid translations of the research instruments and intervention), (ii) an acquired brain injury or substance misuse judged to be the acute cause of the psychotic experiences.

### Additional outcome measures

#### Experiences of psychosis

This was measured by the clinician administered Positive and Negative Syndrome Scale (PANSS) but only the positive subscale was used (Kay & Opler, 1987). Participants can score from 1 (absent) to 7 (extreme) on each item.

#### Depression

Depression was measured by the self-report Beck Depression Inventory brief 7-item measure (BDI-7) (Beck et al., 1997). Participants can score from 0 (not present) to 3 (indicating severe) and have a total score of 21.

#### Hopelessness

The short-form of the Beck Hopelessness Scale (BHS), which is a recently developed 9-item version of the scale validated for psychiatric inpatients, was used to measure hopelessness (Beck & Steer, 1988). Participants can score 0 (not present) or 1 (present) on each item.

#### Personal recovery

This was measured by the 15-item Process of Recovery Questionnaire (QPR) (Law et al., 2014). Participants can score from 0 (disagree strongly) to 4 (agree strongly) on each item.

#### Anxiety

Anxiety was measured on the Generalised Anxiety Disorder-7 item (GAD-7) measure (Spitzer et al., 2006). Participants can score from 0 (not at all) to 3 (nearly every day) on each item. A potential total score can range from 0 to 21.

#### Quality of life

This was measured using the Recovering Quality of Life (REQOL-10) (Keetharuth et al., 2018). Participants rate their quality of life on 10 items from 0 to 4.

#### Functioning

This was assessed on the Global Assessment of Functioning (GAF) measure. GAF scores range from 1 (in some danger of hurting self or others) to 100 (absent or minimal symptoms), examining both symptoms and functioning (Pederson et al., 2020).

#### Mental health and risk

This was measured using the Threshold Assessment Grid (TAG). It has seven domains measuring three areas of safety, risk and needs and disabilities (Slade et al., 2000). Participants can score from 0 (none) to 4 (very severe) on each domain.

### Procedure

This study was carried out in three stages of assessment. Stage one baseline assessments involved participants (n = 80) completing the CRISP alongside all other measures. Data from stage 1 was used to carry out the factor analysis, internal consistency, inter-rater reliability, and concurrent validity of the CRISP. The second stage follow-up involved all n=80 participants being contacted to complete the CRISP again at a 1-week time point to examine for test-retest reliability. Stage 3 (at 2-months) included CRISP trial participants (n=52) being contact to complete all measures (CRISP and all other measures) to examine for sensitivity to change. Due to drop outs, only n=66 (83%) participants completed the stage 2 follow-up and n=30 (58%) completed the stage 3 follow-up.

### Data analysis

All data analysis was conducted using IBM SPSS version 29 (IBM Corp, 2015). Where whole outcome measures were missing, data would be excluded pairwise for the respective analysis. Where less than 25% of individual items were missing from outcome measures, these would be replaced with the measure mean. Data was initially checked for normality through examination of skewness and kurtosis and found to be normally distributed (Kim, 2013). Initially, individual CRISP items were compared using the Pearson’s correlation coefficient to ensure that no items were either extremely highly or poorly correlated. All CRISP items were entered into an exploratory Principal components Analysis (PCA) with Direct Oblimin rotation, and internal consistency was examined for the identified factor’s items. Test-retest reliability was tested for by examining the Pearson correlation coefficients between the CRISP total scores at stage 1 and 2. The CRISP was compared to the PANSS (Kay & Opler, 1987) to examine for criterion validity using Pearson’s correlation analysis. The PANSS was chosen because it is the gold standard measure of symptoms in psychosis. Construct validity was examined through comparisons of the CRISP to all other measures using Pearson’s correlation analysis. Sensitivity to change was calculated by comparing the change score (stage 1 mean score minus the stage 3 mean score) of the CRISP to change score of all other measures. Floor and ceiling effects were determined as present if more than 15% of the sample scored the minimum (0) or maximum (40) score on the SIMS (Terwee et al., 2007).

## Results

A total of 80 participants took part in the study. Demographics can be seen in table 2.

### Initial data scrutiny

Descriptive statistics for the CRISP measure (the means of each individual items, standard deviations, range, skewness, kurtosis and score frequency) can be found in table 3. Individual items from the CRISP were initially screened for their relationship with one another (table 4). If items were either high or low item correlations (<0.2 or >0.9) with more than one other item they would be removed. Items 3 (*I have control over my mental health/experiences of psychosis*), 11 (*I am concerned about housing problems), 12 (I am concerned about financial difficulties*), 14 (*I have strong feelings of anger and frustration*), 12 *(I feel confident that I can look after myself and do day to day tasks if I needed to (e*.*g. Cooking, cleaning, self-care)*), 16 (*I feel I have meaningful and trusting relationships with others (e*.*g. with family, friends, peers))*, 17 (*I am able to ask for help from others when I need it (e*.*g. family, friends, staff))*, 18 (*I feel able to live my life in line with my morals and values*), 19 (*I understand the things I need to do to improve my mental health/experiences of psychosis*) were removed. All remaining 14 items were included in the factor analysis.

### Examination of reliability

*Principal Components analysis with varimax rotation:* The examination of the screen plot (figure 1) and eigenvalues led to 3 factor being identified. The factors explained 64.29% of the variance. Eigen values, variance explained and factor loadings are shown in table 5. All items loaded onto the three factors; “cognitive and emotional symptoms”, “psychosis and risk” and “personal recovery. A final 14-item measure was used for the subsequent analysis, which can be found in the supplementary material.

#### Internal consistency

Internal consistency was examined using the Cronbach alpha statistic. The CRISP showed excellent/good internal consistency with a Cronbach alpha of α= 0.912. Intraclass correlations using a two way mixed model were also examined to test for reliability. The measure had an excellent ICC of 0.912 (CI: 0.880 - 0.938, p <0.001).

#### Reproducibility (test-retest reliability)

The total mean score for the CRISP at stage1 was 29.23 (SD: 11.80) and 29.88 (SD: 10.04) at stage 2. The CRISP showed good test retest reliability with a significant ICC of 0.842 (CI: 0.742 to 0.904,p<0.001).

### Examination of validity

Descriptive statistics and Pearson correlation coefficients for all measures are shown in table 6.

#### Criterion validity

The CRISP illustrated a strong positive correlation with the PANSS, which is considered the gold standard measure of psychosis symptoms measures.

#### Construct validity

The CRISP was highly correlated with many of the other measures (table 7) excluding the TAG and GAF. The TAG and GAF did not correlate with any of the other measures except for a moderate correlation with the PANSS.

#### Sensitivity to change

The mean change and correlation coefficients between time 1 and 3 can be found in table 8. The CRISP was found to have significant correlations with most measures excluding the QPR (−0.340, p=0.082), TAG (0.368, p=0.054) and GAF (−0.190, p-0.332) but neared significant for former two measures. The QPR, TAG and GAF measures also least correlated with the other items.

#### Floor and ceiling effects

The CRISP did not illustrate any floor or ceiling effects. Four (5%) of participants endorsed the lowest possible total score and no participants (0%) endorsed the highest possible score. The frequency of total score endorsements can be found in the supplementary material.

## Discussion

The aim of this study was to develop and examine the psychometric properties of the CRISP. Overall, analysis demonstrated that a 14-item CRISP measure is a reliable and valid tool to assess crisis recovery in patients experiencing psychosis. The CRISP is the first measure of crisis recovery specifically developed for people who experience psychosis and provides a useful step in assessing meaningful outcomes with this population in times of crisis. The outcome measure demonstrated excellent reliability, including both internal consistency and interrater reliability, as well as good overall validity. While it did not show significant correlations with the TAG (Slade et al., 2000) and GAF (Pederson et al., 2020) scales when examined for criterion validity and sensitivity to change, these measures also showed weak correlations with other instruments and exhibited lower internal consistency. This suggests that the TAG (Slade et al., 2000) and GAF (Pederson et al., 2020) may be less reliable and valid in this context.

One of the strengths of the CRISP is its development in partnership with people with lived experience, family/carers and clinicians, which will ensure it measuring relevant factors to recovery from a psychotic crisis (Wood et al., 2023). The measure is also simple and quick to administer, which given the acute and complex nature of the population, means that meaningful outcomes can be examined quickly and with minimal burden with inpatients. Previous research has highlighted the challenges of following up with participants in clinical trials conducted in crisis settings and has recommended the use of simplified and brief outcome measures (Jacobsen et al., 2022), which the CRISIS measure is. The measure is also one of few measures specifically designed and psychometrically tested with inpatient populations. These populations are historically neglected and outcome measures are not often developed specifically for this population (Jacobsen et al., 2024). Researchers examining inpatient populations often utilise outcome measures not specifically developed or tested with this population meaning that there is an increased likelihood of type 1 or type 2 errors. This research now ensure that a psychometrically robust outcome measure is available for researchers conducting research with this population.

This outcome measure also aligns with a recent study which undertook rigorous consensus work to identify key outcome that are important to examine when undertaking psychological intervention research in inpatient settings (Jacobsen et al., 2024). This research identified that the ability to Cope, Hopefulness, Quality of Life, Psychosis Symptoms, Mood, and Self-Harm Behaviours are important key outcomes and all are capture in the CRISP. This means that all key outcomes can be examined in a single measure.

There are limitations to this research. Part of the data used in this paper has been taken from a randomised clinical trial of an adapted CBTp intervention for psychosis in inpatient settings (Wood et al., 2022). This means that some participants (approximately a third) were undertaking a psychological intervention, which would have undoubtedly impacted their scores on their measures, which would have particularly impacted the sensitivity to change analysis. Nevertheless, we were able to still identify that the CRISP had good sensitivity to change despite this. Another limitation was the sample size. Although a reasonable and appropriate size for the type of analysis undertaken, a larger sample size would have provided more robust evidence of the measures psychometric properties.

Another limitation was that we were not able to provide guidance on interpretability or population-specific ratings of measurement as recommended by Terwee et al (2007). We have only been able to administer the measure to a small clinical population of inpatients with psychosis and further research is required to determine these. A final limitation was how the GAF and TAG were administered. The study depended on input from Clinical Studies Officers, which are researchers funded centrally by the Clinical Research Network to support NIHR funded trials recruiting across NHS trusts in the UK. This meant that we had over eight staff members working on the study throughout its duration, which made inter-rater reliability of particularly the TAG and GAF (researcher administered) a particular concern. The researchers had extensive training administering the PANSS, which made this measure administration less concerning, but the TAG and GAF were new measures. We were also not able to calculate inter rater reliability on research administered outcome measures due to the volume of research assistants and lack of study resource which may have reduce measurement consistency and validity.

In conclusion, the CRISP is a reliable and valid outcome measure of psychotic crisis recovery that has been developed with involvement of people with lived experience, family/carers and clinicians. It is a relatively brief measure that examines core outcomes relevant to inpatient and crisis settings.

## Data Availability

All data produced in the present study are available upon reasonable request to the authors

